# Estimating prevalence of subjective cognitive decline in and across international cohort studies of aging: A COSMIC study

**DOI:** 10.1101/2020.05.20.20106526

**Authors:** Susanne Röhr, Alexander Pabst, Steffi G. Riedel-Heller, Frank Jessen, Yuda Turana, Yvonne S. Handajani, Carol Brayne, Fiona E. Matthews, Blossom C.M. Stephan, Richard B. Lipton, Mindy J. Katz, Cuiling Wang, Maëlenn Guerchet, Pierre-Marie Preux, Pascal Mbelesso, Karen Ritchie, Marie-Laure Ancelin, Isabelle Carrière, Antonio Guaita, Annalisa Davin, Roberta Vaccaro, Ki Woong Kim, Ji Won Han, Seung Wan Suh, Suzana Shahar, Normah C. Din, Divya Vanoh, Martin van Boxtel, Sebastian Köhler, Mary Ganguli, Erin P. Jacobsen, Beth E. Snitz, Kaarin J. Anstey, Nicolas Cherbuin, Shuzo Kumagai, Sanmei Chen, Kenji Narazaki, Tze Pin Ng, Qi Gao, Xin Yi Gwee, Henry Brodaty, Nicole A. Kochan, Julian Trollor, Antonio Lobo, Raúl López-Antón, Javier Santabárbara, John D. Crawford, Darren M. Lipnicki, Perminder S. Sachdev, for Cohort Studies of Memory in an International Consortium (COSMIC)

## Abstract

Subjective cognitive decline (SCD) is recognized as a risk stage for Alzheimer’s disease (AD) and other dementias, but its prevalence is not well known. We aimed to use uniform criteria to better estimate SCD prevalence across international cohorts. Therefore, we combined individual participant data for 16 cohorts from 15 countries (members of the COSMIC consortium) and used qualitative and quantitative (Item Response Theory/IRT) harmonization techniques to estimate SCD prevalence. The sample comprised 39,387 cognitively unimpaired individuals above age 60. The prevalence of SCD across studies was around one quarter with both qualitative harmonization/QH (23.8%, 95%CI = 23.3%-24.4%) and IRT (25.6%, 95%CI = 25.1%-26.1%); however, prevalence estimates varied largely between studies (QH: 6.1%, 95%CI = 5.1%-7.0%, to 52.7%, 95%CI = 47.4%-58.0%; IRT: 7.8%, 95%%CI = 6.8%-8.9%, to 52.7%, 95%CI = 47.4%-58.0%). Across studies, SCD prevalence was higher in men than women, in lower levels of education, in Asian and Black African people compared to White people, in lower- and middle-income countries compared to high-income countries, and in studies conducted in later decades. Data harmonization and application of uniform criteria across diverse cohorts yielded more accurate estimates of SCD prevalence. Having a quarter of older individuals with SCD warrants further investigation of its significance, as a risk stage for AD and other dementias, and of ways to help individuals with SCD who seek medical advice.

## Introduction

In light of the projected increase of people living with dementia all around the world, there is a strong interest in early risk stages that may allow early intervention or prevention of dementia. Subjective cognitive decline (SCD) has recently attracted renewed attention on the assumption that it could be the first notable manifestation in the preclinical stage of Alzheimer’s disease (AD) and other dementias [1]. SCD refers to a self-experienced decline in cognitive ability in comparison with a previously normal status and without objective cognitive impairment [2]. The updated AD research framework of the National Institute on Aging and Alzheimer’s Association (NIA-AA) now recognizes SCD within the cognitively unimpaired stage on the cognitive continuum [3]. Thus, SCD is considered a risk stage for dementia. This is supported by evidence from longitudinal epidemiological data that show an increased risk for mild cognitive impairment (MIC) and dementia in individuals with SCD [1].

Importantly, the subjective perception of declining cognitive capacity can also emerge due to conditions other than AD, for example as part of normal aging, MCI, and in association with depression and anxiety [2]. Primarily, SCD is considered a symptom of preclinical AD only in association with AD biomarkers; however, another view is to consider SCD as a broader behavioral phenotype [4] above and beyond preclinical AD that defines a group of people being concerned about their brain health. This is reflected in an increasing number of individuals who seek medical advice because of SCD [5].

Despite the growing research interest in SCD, the concept still faces methodological challenges regarding its operationalization [6]. Historically, the field has lacked a common terminology and definition since the initial description of a “forgetfulness phase” by Reisberg et al. in 1982 [7]. This resulted in the dissemination of a variety of terms, e.g. subjective memory complaints, subjective memory impairment, forgetfulness, subjective cognitive impairment, or cognitive concerns – to name a few. This hurdle has recently been cleared through the introduction of a consensus definition of SCD in preclinical AD by the working group of the Subjective Cognitive Decline Initiative (SCD-I) [2]. A standard approach to measure SCD, however, is still lacking [8]. In light of the evolution of the concept, it is unsurprising that previous findings on SCD epidemiology varied, including prevalence estimates. Early studies investigating more general memory complaints reported prevalence between 22% and 56% in community-based samples [9]. Few studies have estimated SCD prevalence building on the SCD-I criteria. In a sample representing the Greek population aged ≥65 years without psychiatric conditions, 28% of the cognitively unimpaired participants reported SCD [10]. In a German sample of cognitively unimpaired individuals aged ≥75 years, SCD prevalence was 54% [11]. In Chinese residents aged 60 to 80 years, SCD prevalence ranged between 14% and 19% [12]. This, together with a lack of standardized SCD assessment and variations in case definitions, explains variance in reported outcomes. Notably, the occurrence of SCD has almost exclusively been studied in high-income countries (HIC), while evidence from low- and middle-income countries (LMIC) is lacking. The SCD-I emphasized the need for harmonized observational studies that can attenuate some of the limitations associated with SCD operationalization [6].

### Study aims

We aimed to estimate SCD prevalence in cognitively unimpaired older individuals by applying uniform SCD criteria to harmonized data from 16 diverse cohort studies of aging. By doing so, we aimed to minimize the influence of both study level and individual level factors, thereby enhancing the generalizability of findings [13]. Moreover, we aimed to examine differences in SCD prevalence according to sociodemographic factors across studies.

## Methods

### Contributing studies and participants

Cross-sectional population-based individual participant data (IPD) were contributed by 16 member studies of the Cohort Studies of Memory in an International Consortium (COSMIC; https://cheba.unsw.edu.au/consortia/cosmic/studies) [14] (Table 1; see Supplemental material Table e-1 for key references). COSMIC brings together international cohort studies of aging to foster cross-cohort analyses on common factors for cognitive decline and dementia. Study-based data were harmonized and pooled. Baseline data were used for all studies except for two that did not assess all variables needed for SCD classification until later waves, i.e. MoVIES provided data for wave 2 (two years after baseline) and PATH for wave 3 (eight years after baseline). The initial sample included individuals aged at least 60 years and without a dementia diagnosis (Table e-2). This was the default option as some studies excluded prevalent dementia cases as per their study design. For the current study, data represented 15 countries from Africa, Asia, Australia, Europe, and North America.

**Table 1.**
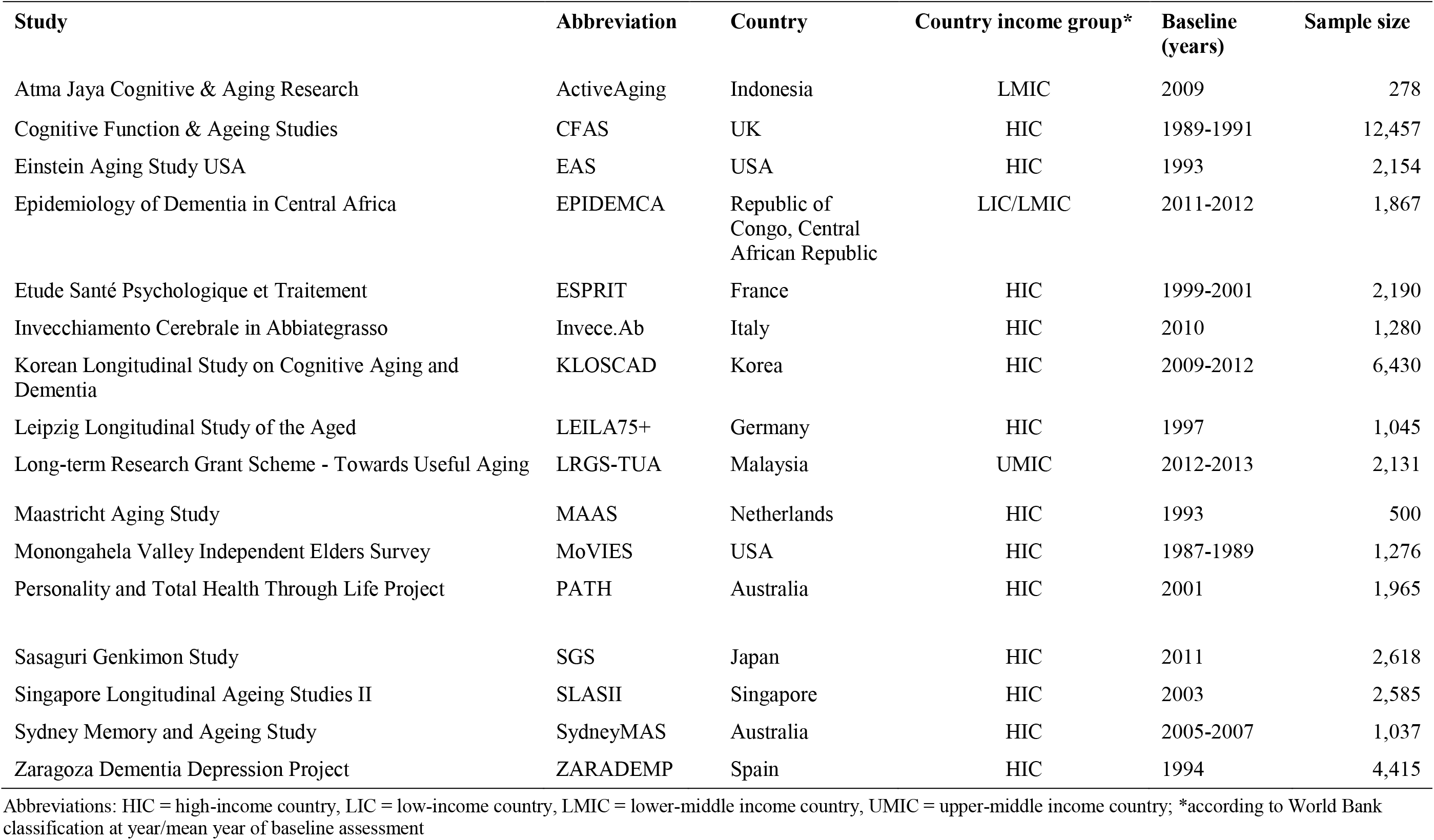
Contributing studies (*n* = 16).

### Ethics

The Human Research Ethics Committee of the University of New South Wales (COSMIC coordinator) approved this study (Ref: HC17292). All contributing studies had previously obtained approval from their respective ethics committees, and it was standard that participants provided written informed consent.

### Demographics

Information included age, gender, education, ethnicity, country income, and decade in which a study was conducted. Education was provided as years for most studies. For four cohorts (ActiveAgeing, EPIDEMCA, ESPRIT, MAAS), education data were provided as categories. For harmonization purposes, categories were assigned discrete year values based on the local education system as informed by the study leaders. A fifth cohort (MoVIES) provided category data for all participants and year values for 73.4%. These data were used to calculate a mean year value for each category that was assigned to individuals missing education year data. For subgroup analysis only, years of education were re-categorized according to the UNESCO International Standard Classification of Education (ISCED) 2011 into pre-/primary education (zero to five years), secondary education (six to nine years), upper-/post-secondary education (ten to 14 years), and tertiary education (>14 years) [15].

Ethnicity was recorded according to self-report. For studies lacking IPD for ethnicity, participants’ ethnicity was assigned as the majority ethnicity of the study sample as informed by the study leaders.

Country income was categorized according to the World Bank classification corresponding to the year or mean year in which baseline assessments took place. It is based on the gross national income per capita, i.e. low-, lower middle-, upper middle-, and high-income country.

To account for a potential impact of time trends regarding dementia awareness, we furthermore considered the decade, in which studies were initiated (<1999, 2000-2009, >2009).

### Assessment of self-experienced decline in cognitive capacity

Assessment of a self-experienced decline in cognitive capacity varied between studies, though answers to all questions were self-reported during face-to-face interviews. Six studies used a single self-developed question, seven studies used a battery of self-composed questions, and one study used the Subjective Memory Complaints Questionnaire (SMCQ) [16]. Additionally, two studies (ActiveAging and LRGS-TUA) provided a relevant item from the Geriatric Depression Scale (GDS): “Do you feel you have more problems with memory than most?” [17].

In PATH, the Informant Questionnaire on Cognitive Decline in the Elderly (IQCODE) was administered as self-report, including seven items that addressed whether different facets of memory and recall had changed over time [18]. Data on all identified indicators were then synthesized and harmonized.

One fundamental challenge of integrative data analysis is the evaluation and statistical consideration of between-sample heterogeneity. Since the majority of contributing studies used multiple items to assess a self-experienced decline in cognitive capacity, we followed two complementary strategies to prepare data.

### Qualitative harmonization of items for a self-experienced decline in cognitive capacity

The first harmonization strategy followed a qualitative approach [19]. Authors SR and AP independently compared all items assessing self-experienced decline in cognitive capacity between studies and then selected and harmonized them by matching semantically similar items; i.e. if more than one item was available. Inter-rater agreement was high *(K* = 0.97). The original scales for all items were transformed, if necessary, to provide binary responses of presence or absence of a self-experienced decline in cognitive capacity. As a result, we identified and harmonized one common item that bridges the measurement of a self-experienced decline in cognitive capacity across studies (we refer to this item as “item 1”). Table e-3 shows the assessment of the selected items underlying the harmonization of item 1 across studies. Commonly, these items broadly addressed whether the study participant had problems or difficulties with memory; MAAS provided a single item assessing cognitive failures. For six studies, only data on this bridging item 1 was available. For the remaining ten studies, information on additional items was available and considered for harmonization. In addition to the bridging item 1, information from studies with further items available was considered. Overall, we were able to identify and harmonize a total of 32 different items for IRT analysis (Table e-4).

### Quantitative harmonization of items for a self-experienced decline in cognitive capacity

The second harmonization strategy followed a quantitative approach in order to develop a measurement model for both common and study-unique items. The goal was to generate scale scores for a generic construct of self-experienced cognitive decline, which is commensurate in meaning and metric across studies and study subpopulations [20]. In particular, we used the 2-Parameter Logistic (2-PL) Item Response Theory (IRT) model as a psychometric approach to evaluate measurement equivalence of items across the 16 studies. IRT allows the localization of both item difficulty and person characteristics on a common latent “trait” that represents self-experienced decline in cognitive capacity, while controlling for between-study heterogeneity of measurement. Higher difficulty values indicate a higher likelihood of a positive response to a certain item [20]. First, we applied an automated bottom-up stepwise item selection procedure to identify a core set of items that builds the basis for IRT analysis [21]. The procedure revealed that 18 of the 32 items (see Table e-4) form a unidimensional factor with acceptable scalability (Loevinger’s *H_ij_* = 0.43) and good reliability (Cronbach’s Alpha = 0.81). Inspection of parameters of the fitted 2-PL IRT model (i.e., difficulty, discrimination), as well as the evaluation of item characteristics curves led to the exclusion of two additional items with insufficient psychometric properties (item 2, item 5), resulting in a final IRT model with 16 items with partial factorial invariance and acceptable model fit (Root Mean Square Error of Approximation =.02; Comparative Fit Index =.87; Tucker Lewis Index =.85; Standardized Root Mean Square Residuals =.09). Finally, we dichotomized the predicted latent score obtained from the final IRT model at *theta* = 0 to differentiate individuals with a higher likelihood of present symptoms from those with lower estimated likelihood [22].

### Cognitive impairment

The Mini-Mental State Examination (MMSE) was used to assess cognitive impairment [23]. This was the default option as the MMSE was assessed in all but two studies, for which MMSE scores could be derived from similar tests. For the EAS, Blessed Information Memory Concentration scores were converted to MMSE scores using a validated formula [24, 25]. For the EPIDEMCA cohort, Community Screening Interview for Dementia scores were converted to MMSE scores using a co-calibration table [26, 27].

### Functional ability

Functional ability was based on the assessment of instrumental activities of daily living (IADL). Nine studies used the Lawton and Brody IADL Scale [28]. Seven studies each used a different scale, though all had large overlap in key activities (e.g., food preparation, shopping). All instruments are listed in Table S5. For all studies, higher scores indicated higher functionality (after reverse scoring for EPIDEMCA and SydneyMAS).

### Depressive and anxiety symptomatology

All studies contributed data for depression and 12 studies for anxiety (anxiety data were not collected in ActiveAging, LEILA75+, LRGS-TUA, and MoVIES). Data were harmonized as per previous COSMIC reports, with all available information considered [29, 30]. Depression was indicated by any of scale scores meeting cut-off, expert diagnosis, self-report, treatment, or use of antidepressant medication (Table e-6). Similarly, anxiety disorders were indicated by any of scale scores meeting cut-off, self-report, or the use of anxiolytics (Table e-7).

### Operationalization of SCD cases

SCD cases were uniformly defined and operationalized according to current criteria for SCD and followed recommendations on the implementation of SCD in research, i.e. endorsement of a self-experienced decline in cognitive capacity in the absence of objective cognitive impairment (criterion 1), unimpaired functional ability (criterion 2), exclusion of major depression (criterion 3), and exclusion of anxiety disorder (criterion 4) [1, 6].

Endorsement of a self-experienced decline in cognitive capacity and the presence or absence of major depression or anxiety disorder were determined as above.

Unimpaired cognitive functioning was operationalized as a score higher than 1.5 SDs below the mean of the study-based age-, gender-, and education-adjusted MMSE score. Likewise, unimpaired functional ability was operationalized as a score higher than 1.5 SDs below the mean of the study-based age-, gender-, education-adjusted IADL scores.

### Data analysis

Prevalence in our study was defined as the number of SCD cases divided by the total number of cognitively unimpaired individuals, and estimates for both the qualitative harmonization (QH) and quantitative IRT harmonization approach are presented as percentages with 95% confidence intervals (95%CI). All estimates were stratified by study and subsequently by demographic subgroups (age, gender, education, ethnicity, country income, decade). Study-specific prevalence estimates as well as subgroup-specific prevalence estimates and corresponding tests regarding differences in education, ethnicity, country income, and decade were adjusted for age and gender, using the total sample of all 16 studies included in the analysis as the standard population. Gender differences were adjusted for age and vice versa. This allowed for the direct comparison of prevalence estimates between studies and subgroups, including different distributions of core sociodemographic variables [31]. Subgroup comparisons of proportions were tested using Pearson’s Chi-square test with Rao/Scott correction. Overall prevalence estimates across studies are reported as unstandardized, since data on standard populations stratified by age and gender were not available.

To illustrate the impact of the individual criteria to quantify SCD cases, we show frequency rates criteria-wise, i.e. cumulatively applying the four criteria for SCD.

Sensitivity analysis for IRT results was performed excluding studies with only the bridging item (‘item 1’) available. Furthermore, we inspected the contingency of the results of the two harmonization strategies (QH and IRT). Analyses were performed in Stata 15 MP (Stata Corp, College Station, TX) and R version 3.5.0 [32].

## RESULTS

### Sample

The initial sample comprised 44,228 dementia-free individuals aged at least 60 years. Inspection of the initial sample led to the exclusion of 1,037 (2.3%) individuals due to study-based missing information on any of the 32 SCD items and 1,049 (2.4%) individuals due to missing information on demographics and/or MMSE. In addition, 2,755 (6.2%) individuals classified as cognitively impaired were excluded, resulting in an analytical sample of 39,387 individuals (Table 2). Mean age of participants in the analytical sample was 73.1 years *(SD* = 7.1; range = 60-105 years). The proportion of women was 57.7%. Education was 9.1 years *(SD* = 4.4) on average.

**Table 2.**
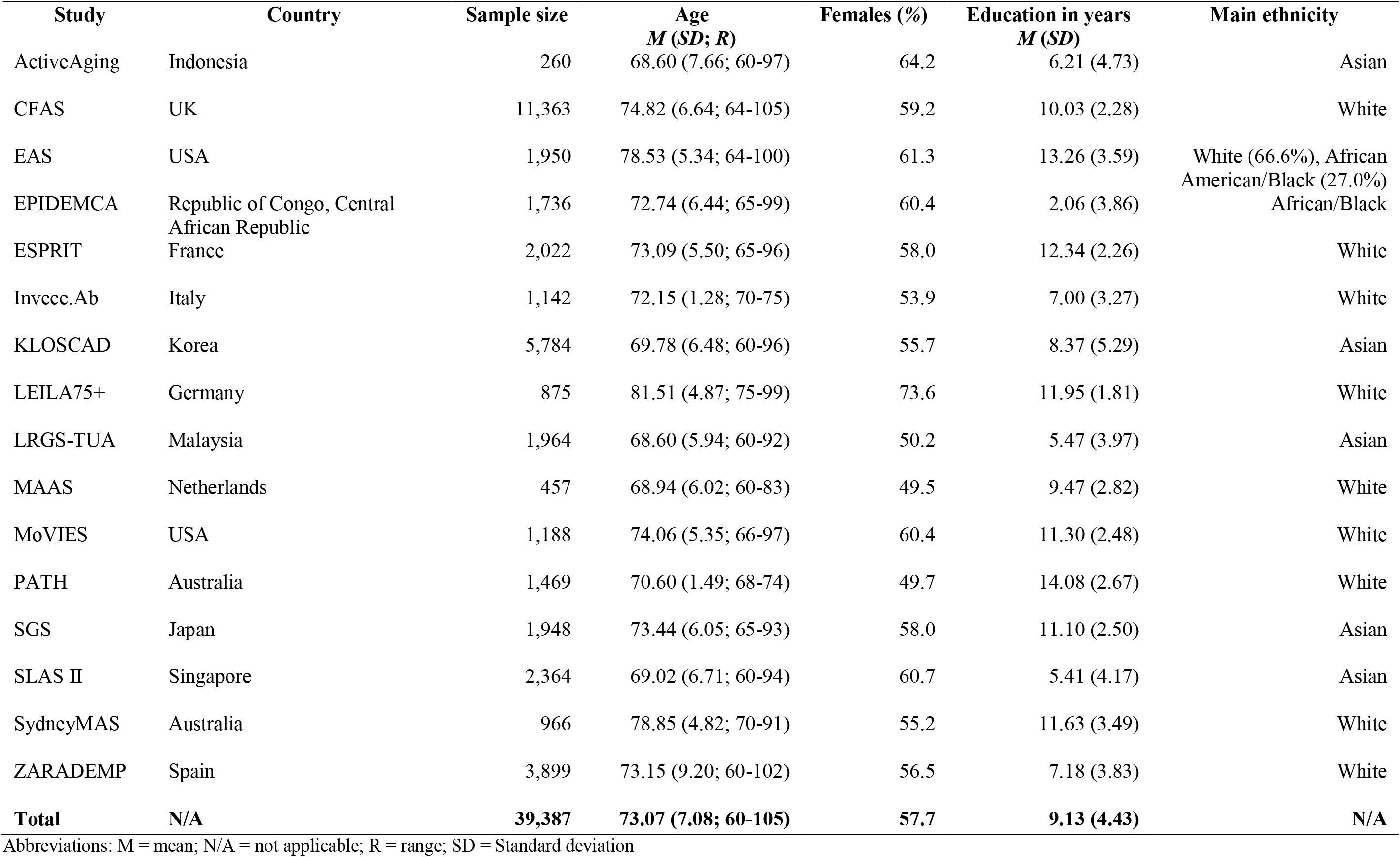
Baseline characteristics of the analytical sample (n = 39,387).

### Prevalence of SCD in and across studies

Overall, both the QH and IRT approach robustly suggested a SCD prevalence of roughly one in four in the older population without cognitive impairment across studies (QH: 23.8%, 95%CI = 23.3%-24.4%; IRT: 25.6%, 95%CI = 25.1%-26.1%). Study-based age- and gender-standardized SCD prevalence estimates varied largely, ranging from 6.1% (95%CI = 5.1%-7.0%) to 52.7% (95%CI = 47.4%-58.0%) for QH and 7.8% (95%CI = 6.8%-8.9%) to 52.7% (95%CI = 47.4%-58.0%) for IRT. All study-based SCD prevalence estimates according to QH and IRT are shown in the last column in Tables 3 and 4, respectively.

**Table 3.** Study-based age- and gender-standardized prevalence estimates for subjective cognitive decline (SCD; last column) and cumulative frequency estimates for the stepwise application of SCD operationalization criteria according to qualitative harmonization.

| Study | SCD operationalization criteria |  |  |  |
| --- | --- | --- | --- | --- |
|  | Criterion 1<br>Endorsement of a self-experienced decline in cognitive capacity without objective cognitive impairment | + Criterion 2<br>Intact functional ability | + Criterion 3<br>No major depression | + Criterion 4<br>No anxiety disorder |
|  | % (95%CI) |  |  |  |
| ActiveAgeing | 27.98 (21.44-34.52) | 25.81 (19.41-32.20) | 25.73 (19.36-32.10) | N/A |
| CFAS | 17.04 (15.45-18.64) | 15.59 (14.00-17.18) | 12.73 (11.41-14.05) | 6.34 (5.57-7.20) |
| EAS | 15.84 (12.57-19.10) | 13.82 (10.90-16.73) | 15.23 (10.95-19.51) | 12.05 (5.80-18.30) |
| EPIDEMCA | 50.52 (48.36-52.69) | 47.74 (45.51-49.98) | 25.83 (23.80-27.86) | 23.86 (21.93-25.79) |
| ESPRIT | 18.17 (16.52-19.82) | 16.60 (15.02-18.18) | 13.36 (11.92-14.80) | 10.35 (9.11-11.60) |
| Invece.Ab | 14.60 (11.63-17.56) | 14.02 (10.89-17.16) | 12.31 (9.43-15.18) | 10.76 (8.28-13.23) |
| KLOSCAD | 63.65 (62.28-65.02) | 58.02 (57.88-59.46) | 42.25 (40.82-43.68) | 42.27 (40.90-43.66) |
| LEILA75+ | 12.88 (11.65-14.12) | 11.70 (10.48-12.92) | 9.80 (8.64-10.96) | N/A |
| LRGS-TUA | 51.23 (48.15-54.31) | 47.39 (44.32-50.47) | 46.84 (43.80-49.87) | N/A |
| MAAS | 65.26 (60.33-70.20) | 60.52 (55.35-65.69) | 54.97 (49.43-60.50) | 52.70 (47.43-57.96) |
| MoVIES | 32.82 (30.35-35.32) | 30.45 (28.01-32.90) | 27.27 (24.89-29.65) | N/A |
| PATH | 8.95 (7.89-10.01) | 8.23 (7.21-9.26) | 7.34 (6.36-8.33) | 6.06 (5.13-6.99) |
| SGS | 18.47 (16.82-20.13) | 16.52 (14.93-18.10) | 10.24 (8.94-11.55) | 9.91 (8.64 (11.18) |
| SLAS II | 12.16 (10.53-13.79) | 10.82 (9.27-12.36) | 10.28 (8.78-11.78) | 9.89 (8.50-11.28) |
| SydneyMAS | 42.77 (40.49-45.04) | 39.10 (36.70-41.51) | 32.58 (30.14-35.01) | 26.98 (24.55-29.40) |
| ZARADEMP | 44.68 (42.98-46.38) | 42.04 (40.31-43.77) | 29.42 (27.77-31.06) | 28.12 (26.50-29.74) |
| **Total\*** | **33.03 (32.56-33.49)** | **30.46 (30.01-30.92)** | **24.08 (23.65-24.51)** | **23.83 (23.29-24.36)** |

Abbreviations: 95% CI = 95% confidence interval; N/A = not available; \*unstandardized

**Table 4.** Study-based age- and gender-standardized prevalence estimates for subjective cognitive decline (SCD; last column) and cumulative frequency estimates for the stepwise application of SCD operationalization criteria according to quantitative harmonization/Item response Theory (IRT) modelling.

| Study | SCD operationalization criteria |  |  |  |
| --- | --- | --- | --- | --- |
|  | Criterion 1<br>Endorsement of a self-experienced decline in cognitive capacity without objective cognitive impairment | + Criterion 2<br>Intact functional ability | + Criterion 3<br>No major depression | + Criterion 4<br>No anxiety disorder |
| Prevalence estimates in % (95%CI) |  |  |  |  |
| ActiveAgeing | 27.97 (21.43-34.51) | 25.80 (19.40-32.20) | 25.73 (19.36-32.10) | N/A |
| CFAS | 23.44 (21.77-25.12) | 21.59 (19.91-23.26) | 17.78 (16.36-19.20) | 9.60 (8.63-10.58) |
| EAS | 50.41 (45.20-55.62) | 46.06 (41.06-51.06) | 38.71 (34.07-43.34) | 25.91 (19.33-32.48) |
| EPIDEMCA | 56.28 (54.18-58.38) | 53.02 (50.83-55.21) | 30.64 (28.51-32.77) | 28.41 (26.38-30.44) |
| ESPRIT | 31.26 (29.34-33.18) | 28.76 (26.89-30.63) | 23.97 (22.20-25.74) | 18.94 (17.36-20.51) |
| Invece.Ab | 14.61 (11.64-17.58) | 14.03 (10.90-17.17) | 12.32 (9.44-15.20) | 10.77 (8.28-13.26) |
| KLOSCAD | 55.51 (54.09-56.92) | 50.55 (49.10-52.00) | 36.01 (34.62-37.40) | 35.97 (34.62-37.31) |
| LEILA75+ | 12.89 (11.66-14.13) | 11.71 (10.49-12.94) | 9.81 (8.65-10.97) | N/A |
| LRGS-TUA | 51.23 (48.15-54.31) | 47.40 (44.32-50.47) | 46.84 (43.80-49.87) | N/A |
| MAAS | 65.26 (60.32-70.20) | 60.52 (55.34-65.69) | 54.97 (49.43-60.51) | 52.70 (47.43-57.98) |
| MoVIES | 29.39 (26.97-31.81) | 27.20 (24.83-29.57) | 23.87 (21.59-26.15) | N/A |
| PATH | 11.68 (10.50-12.86) | 10.76 (9.62-11.90) | 9.63 (8.54-10.73) | 7.83 (6.80-8.87) |
| SGS | 18.47 (16.81-20.12) | 16.51 (14.93-18.09) | 10.24 (8.93-11.54) | 9.91 (8.64-11.18) |
| SLAS II | 25.48 (23.37-27.59) | 22.87 (20.83-24.91) | 22.17 (20.16-24.18) | 21.18 (19.31-23.05) |
| SydneyMAS | 45.81 (43.41-46.39) | 42.72 (40.48-44.95) | 35.87 (33.55-38.19) | 29.88 (27.54-32.22) |
| ZARADEMP | 44.69 (42.99-46.39) | 42.04 (40.32-43.77) | 30.64 (28.51-32.77) | 28.12 (26.50-29.74) |
| **Total\*** | **37.32 (36.85-37.79)** | **34.49 (34.02-34.97)** | **27.75 (27.31-28.20)** | **25.60 (25.06-26.14)** |

Abbreviations: 95% CI = 95% confidence interval; N/A = not available; \*unstandardized

### Proportions according to cumulative criteria application

The columns of Tables 3 and 4 show the proportions of the incremental application of the four operationalization criteria for SCD. There was a particular difference in proportions between functional criteria (endorsement of a self-experienced decline in cognitive capacity without objective cognitive impairment and functional ability) and mood criteria (depressive and anxiety symptomatology). When in addition to functional criteria depressive and anxiety symptomatology were addressed, overall proportions decreased from 30.5% (95%CI = 30.0%-30.9%) to 23.8% (95%CI = 23.3%-24.4%) in QH and from 34.5% (95%CI = 34.0%-35.0%) to 25.6% (95%CI = 25.1%-26.1%) in IRT.

### Prevalence of SCD according to age group and gender across studies

Reported subgroup results are based on IRT analysis, applying all four SCD criteria. Gender differences were adjusted for age and vice versa. Overall, SCD prevalence was somewhat higher in men compared to women (26.6% vs. 24.9%; *χ*^2^(1) = 8.54; *p* =.003). SCD prevalence significantly differed according to age group (*χ*^2^(5) = 51.31; *p* <.001), which also applied when stratified for gender (men: *χ*^2^(5) = 32.42; *p* <.001; women: *χ*^2^(5) = 29.81; *p* <.001). SCD prevalence was 27.4% (95%CI = 25.8%-28.9%) in ages 60-64 years, then decreased to 23.2% (95% CI = 22.1%-24.2%) in ages 65-69 years, thereafter increased to 24.5% (95%CI = 23.6%-25.5%) in ages 70-74 years, 27.9 (95%CI = 26.5%-29.3%) in ages 75-79 years, and 28.1% (95%CI = 26.2%-30.0%) in ages 80-84 years and 28.1% (95%CI = 25.7%-30.9%) in ages 85+ years. Overall, there was no clear pattern associated with age, except for those aged 65-74 years generally having a lower prevalence than all other age groups. Figure 1 additionally shows SCD prevalence according to age groups stratified by gender.

**Figure 1.**
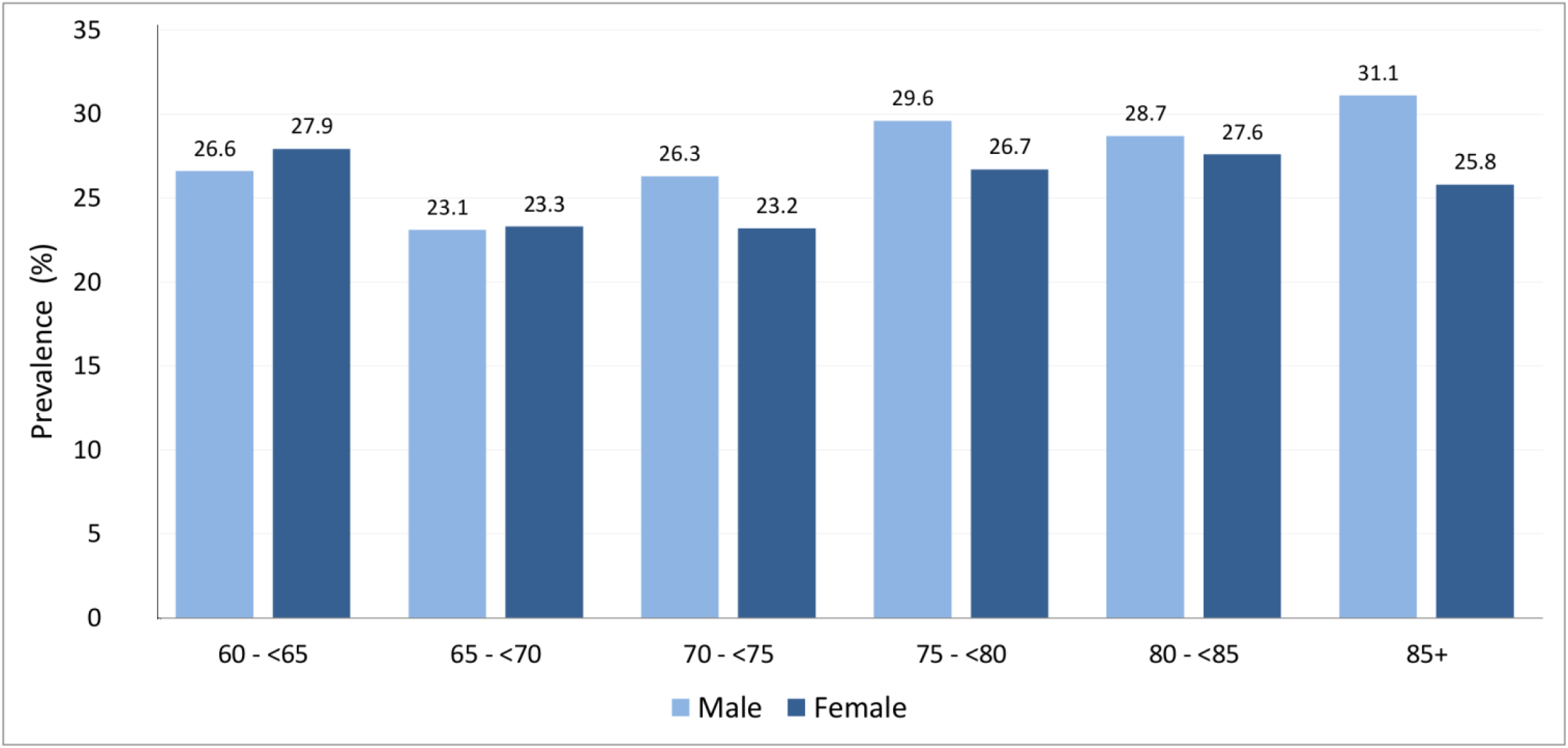
SCD prevalence according to age groups and gender in older individuals (≥ 60 years) without cognitive impairment across international cohort studies (percentages). Estimates are based on Item response Theory (IRT) analysis.

#### Prevalence of SCD according to education across studies

Age- and gender-standardized SCD prevalence significantly differed according to level of education (*χ*^2^(3) = 68.37; *p* <.001). Individuals with pre-/primary education had higher prevalence of SCD (29.0%; 95%CI = 27.9%-30.2%) compared to individuals with secondary education (25.0%; 95%CI = 24.1%-25.9%), secondary upper/post education (22.7%; 95%CI = 21.8%-23.8%), and tertiary education (26.4%; 95%CI = 24.8%-28.0%).

#### Prevalence of SCD according to ethnicity across studies

Due to missing information on self-reported ethnicity, this subgroup analysis was based on a subsample of n = 23,641 individuals. Age- and gender-standardized SCD prevalence in White people (24.3%; 95%CI = 23.5%-25.1%) was significantly lower than in Asian people (27.2%; 95%CI = 26.3%-28.1%) and Black African people (28.2%; 95%CI = 26.2%-30.3%; *χ*^2^(2) = 29.39;*p* <.001). Data availability did not allow including further ethnic groups.

#### Prevalence of SCD according to country income across studies

Age- and gender-standardized SCD prevalence was significantly associated with country income (*χ*^2^(2) = 9.56; *p* <.001). The lower the country income, the higher the SCD prevalence, increasing from 25.1% (95%CI = 24.5%-25.6%) in HIC to 27.4% (95%CI = 24.6%-30.5%) in LMIC to 29.3% (95%CI = 26.4%-32.3%) in LIC.

#### Prevalence of SCD according to decade of study conduction

Age- and gender-standardized SCD prevalence tended to differ by decade (*χ*^2^(2) = 47.77; *p* <.001), showing an increase over time from 22.4% (95%CI = 21.4%-23.3%) before 1999, then 24.5% (95%CI = 23.1%-25.9%) in the years 2000-2009 to 26.3% (95%CI = 25.9%-27.4%) after 2009.

#### Sensitivity analysis

Results of the IRT based sensitivity analyses of only studies that used multiple items to assess a subjective experience of cognitive decline are shown in Table e-8, further supporting a pooled SCD prevalence of one in four (26.1%; 95%CI = 25.4%-26.7%).

The contingency of the results returned by the two harmonization approaches was continuously high across prevalence estimates. Using all four operationalization criteria, SCD case classification based on “item 1” by QH and IRT corresponded in 91.4% (*Φ* = 0.77) and in studies with multiple items in 88.0% (*Φ* = 0.68) of all cases vs. non-cases.

### Discussion

We estimated the prevalence of subjective cognitive decline (SCD) by applying uniform criteria to harmonized individual participant data (IPD). Data represented 16 international population-based cohort studies from 15 countries with over 39,000 individuals at least 60 years old. Across studies, qualitative harmonization (QH) and quantitative harmonization using Item Response Theory (IRT) both robustly suggested a SCD prevalence of roughly one quarter (23.8% and 25.6%) in cognitively unimpaired older individuals. Still, prevalence estimates varied largely between studies (QH: 6.1%-52.7%; IRT: 7.8%-52.7%). However, applying uniform criteria for SCD operationalization to harmonized data helps to reduce the degree of variation in study-based estimates. This is a strength of our study as SCD assessment greatly differed between cohorts and different measurements are known to be associated with outcome variance [33]. Indeed, differences in prevalence estimates are directly associated with test accuracy, and the majority of the items the cohorts used to measure self-experienced decline in cognitive capacity were not psychometrically evaluated. Used in isolation, these potentially imperfect items may give false positive or false negative results, resulting in biased prevalence estimates. However, using multiple items and multiple population groups helps to increase psychometric accuracy, and thus a more accurate estimate of prevalence across studies can be neared [34]. Additionally, IRT analysis allowed heterogeneity between studies to be addressed statistically, by selecting a set of items that form a unidimensional scale, thus giving confidence that all items measured one construct. Hence, our study suggests that the prevalence of SCD in cognitively unimpaired older individuals is rather banded around 25%, which may give a more accurate idea of SCD prevalence than estimates of single studies do, which are potentially more heavily influenced by measurement issues.

Our results support the notion that SCD is frequent in old age. It is clear that only a proportion of these cases reflect pathological change due to AD or other dementias. The list of non-neurodegenerative conditions that can lead to subjective decline in cognition is long, including, apart from major depression and anxiety, normal aging, various psychiatric, neurologic and medical disorders, substance abuse and medication, as well as personality and cultural factors [2]. The current SCD research definition cannot rule out all other causes, and this is likely reflected in the range of prevalence estimates we found. In this regard, it is interesting to note the difference it made when major depression and anxiety disorders were considered as exclusion criteria in addition to cognitive impairment and functional inability (which some studies have only used), reducing estimates from roughly one third to one quarter. While the exclusion of depression and anxiety disorder is accepted in the criteria, it does highlight the impact criteria can have on prevalence estimates. In addition, many of the above named conditions are associated with an increased risk for cognitive decline and dementia themselves. Vice versa, specific symptoms, especially depressive and anxiety symptoms, can also be a consequence of neuropathological change or co-occur, so that the relationship between SCD, these conditions and neurodegeneration is more complex [2]. In this regard, it seems useful to explore SCD in conjunction with other behavioral symptoms [35]. Like SCD, problems with mood, anxiety, drive, perception, sleep, appetite, agitation, and aggression can be precursors to cognitive decline and dementia, as summarized in the construct mild behavioral impairment (MBI) [36]. Future investigations could link the two behavioral concepts rather than studying them independently.

Age- and gender-standardized SCD prevalence differed regarding sociodemographic factors. We found slightly higher SCD prevalence in men compared to women. The literature on gender differences in SCD prevalence is inconsistent, some reporting higher prevalence in women [37, 38], others in men [39] or no difference [40]. For both men and women, SCD prevalence differed by age; however, without a clear pattern. This is similar to MCI prevalence, but different to dementia prevalence that shows an increase with aging [41].

SCD prevalence was highest in individuals with less education, as previously reported by others [42]. While the prevalence decreased with increasing levels of education, there was a tendency to increase again among those with the highest education levels. In general, higher educational attainment is thought to provide resilience against neuropathology [43]. This could be expected to lead to a delay in symptom onset and therefore, potentially, a lower SCD prevalence associated with higher levels of education. However, our finding of higher SCD prevalence in those with the highest education suggests otherwise, and possibly points to increased awareness of or alertness to subtle cognitive changes in this group.

Regarding ethnicity, we found higher SCD prevalence in individuals of Asian and African ancestry compared to individuals of European ancestry. SCD and ethnicity is largely unexplored. One study within the US reported similar levels of SCD for African Americans and Whites [44], whereas in another SCD was lower in Asians compared to Whites and highest in Black Americans and American Indians [42]. Our results should be interpreted with caution as data on ethnicity were limited and may not be representative. Additionally, SCD prevalence was lower in HIC compared to LMIC. From an ecobiopsychosocial perspective, this supports the notion that environments with higher socioeconomic resources (e.g., better health care, better educational opportunities, better lifestyle infrastructure) may be beneficial for population health [45]. Indeed, dementia incidence has been observed to have slightly declined in recent decades in Western high-income countries, supporting such an assumption [46]. Opposed to that, we found a trend of increasing SCD prevalence over decades from before 1999 to after 2009. On the one hand, this could be attributed to increasing public health awareness regarding dementia, or, on the other hand, to the fact, that latter data included more studies from LMIC whereas early studies were exclusively from HIC. However, as much of the increase in numbers of people living with dementia takes place in LMIC [47], there may be indeed a trend towards increasing SCD.

Regardless of causes or consequences, SCD is a serious issue for individuals who experience it. SCD has been associated with concerns [48], lower health-related quality of life [49], increased help-seeking behavior and health care utilization [50]. Thus, SCD has a negative impact on the individual, but also on society through creating additional costs [50]. Having roughly one quarter of the cognitively unimpaired older population experiencing SCD poses the question ‘what to do about it?’. A systematic review and meta-analysis of interventions for SCD targeting well-being, meta-cognition and/or objective cognitive performance reported a lack of high-quality research, but nevertheless found that psychological interventions may be beneficial for well-being and meta-cognition in SCD, though not for cognitive performance [51]. Furthermore, the same study reported a lack of evidence regarding lifestyle and pharmacological interventions. The SCD-I recently argued in support of tailored diagnostic processes that identify underlying medical conditions in individuals with SCD who present to physicians [5]. If no cause can be identified, they suggest to inform about SCD and dementia risk. From there, a watch and wait strategy could be adopted. A comprehensive approach to deal with SCD, if no treatable underlying condition can identified, is perhaps education about modifiable health and lifestyle factors for brain health. Increasing evidence highlights that, among other factors, improved management of diabetes, hypertension and obesity as well as proactive lifestyle behaviors regarding physical, cognitive and social activity, can promote brain health and may mitigate dementia risk [52]. It could be beneficial to adopt such strategies as early as in SCD.

### Limitations

We were able to consider a set of the most agreed upon criteria to define and operationalize SCD; however, there may be factors not considered in these criteria that influence prevalence estimates of SCD. From this perspective, our reported SCD prevalence may be an overestimate.

Regarding IRT-based estimation of prevalence, we were not able to specify a cutoff other than *theta* = 0 to differentiate between SCD cases and non-cases, as a gold standard is not available. Moreover, fit indices of our IRT models were acceptable, but indicated room for improvement, which should be considered when possibly developing a standardized measurement of SCD. Though we were able to methodologically tackle heterogeneity in SCD measurement across studies, estimates for individual studies are likely influenced by the type of questions asked. This could be one of the reasons for the large differences in prevalence between the studies, and calls for a standardized and psychometrically sound instrument for SCD. Also, many items to assess a self-experience in cognitive capacity did not cover perceptions of change over time, the majority instead asked about current problems with memory – an acknowledged limitation in SCD research. Future studies may also explore how study-based characteristics beyond age and gender contribute to SCD prevalence variation between studies. Ultimately, only standardized and valid SCD measurement will overcome these limitations, and the SCD field should focus on the development of such an instrument. Our IRT-based item analysis can provide useful information for this. For now, the comparable results from two complimentary approaches to estimate SCD prevalence across a set of diverse studies, further supported by similar results from sensitivity analyses, provide a more accurate picture of SCD occurrence.

### Conclusions

One in four cognitively unimpaired individuals above 60 years of age is estimated to experience and report SCD. However, SCD is likely to indicate preclinical AD or other dementing disorders in only the minority of cases. Nevertheless, the frequent occurrence of SCD warrants further research of its significance in preclinical AD and non-AD dementias, and, importantly, on ways to manage SCD in clinical practice. The development and application of a standardized measure to assess SCD is imperative to further our understanding of SCD.

## Data Availability

Data and material in relation to the manuscript are available for researchers from the corresponding author upon reasonable request.

## Declarations

### Ethics approval

This study was performed in line with the principles of the Declaration of Helsinki. The Human Research Ethics Committee of the University of New South Wales (COSMIC coordinator) approved this study (Ref: HC17292). All contributing studies had previously obtained approval from their respective ethics committees (see below). Informed consent was obtained from all individual participants by the contributing studies.

| **Study** | **Institutional Review Board** |
| --- | --- |
| ActiveAging | School of Medicine and Health Sciences Atma Jaya Catholic University of Indonesia Ethics Committee (approval 03/04/3014&01/05/2015) |
| CFAS | Anglia and Oxford Multi-centre Research Ethics Committee (MREC) - 99/5/22; Eastern MREC – 99/5/22; Eastern MREC – 05/MRE05/37; NRES Committee East of England – 05/MRE05/37 |
| EAS | Albert Einstein College of Medicine Institutional Review Board (Approval#1996-175) |
| EPIDEMCA | Ethical committees, supervised by Ministry of Public Health in CAR (8/UB/FACSS/CSCVPER/11) and the Comité d’Ethique de la Recherche en Sciences de Santé in ROC (00000204/DGRST/CERSSA), approved the study protocol, as well as the “Comité de la Protection des Personnes Sud-Ouest Outre-Mer” in France (SOOM4/CE/3). |
| ESPRIT | Ethics committee (CCPPRB) of the Kremlin Bicetre hospital (n° registered 99-28) |
| Invece.Ab | Ethics Committee of the University of Pavia (#3/2009) |
| KLOSCAD | Institutional Review Board of Seoul National University Bundang Hospital, Korea (IRB No. B-0912/089-010) |
| LEILA75+ | Ethics committee of the University of Leipzig (C7 79934700) |
| LRGS-TUA | Medical Research and Ethics Committee of Universiti Kebangsaan Malaysia (NN-060-2013).<br>Ministry of Education Longterm Research Grant Scheme (LRGS/BU/2012/UKM–UKM/K/01). |
| MAAS | Ethics committee of Maastricht University Medical Centre (MEC05-107) |
| MoVIES | University of Pittsburgh Institutional Review Board (IRB# 961263-0110) |
| PATH | Australian National University Human Research Ethics Committee (#M9807, #2002/189, #2006/314, # 2010/542, #2001/2, #2009/039) |
| SGS | Institutional Review Board of the Institute of Health Science, Kyushu University (IHS-2010-22) |
| SLASII | National University of Singapore Institutional Review Board (NUS IRB 04-140, 17 June 2004) |
| Sydney MAS | University of New South Wales Human Research Ethics Committee (approval #14327) |
| ZARADEMP | Ethics committee of the Zaragoza University Hospital (CEICA # CP16/2012) |

